# Variability in Health Literacy and Learning Preferences on Peripheral Arterial Disease Among Chinese-speaking Communities in California

**DOI:** 10.64898/2026.06.29.26356871

**Authors:** Chia-Ding Shih, Patcharathorn Pookun, Benjamin Zhang, Jenny Yuan, Kayla Lim, Kossi M. Senagbe, Emily R. Rosario, Tze-Woei Tan

**Affiliations:** Division of Vascular Surgery and Endovascular Therapy, Keck Medicine of USC, Los Angeles, CA, USA; Samuel Merritt University College of Podiatric Medicine, Oakland, CA, USA; Research Institute, Casa Colina Hospital and Centers for Healthcare, Pomona, CA, USA

**Keywords:** Health literacy, community health, population health, peripheral arterial disease

## Abstract

**BACKGROUND:** Peripheral Artery Disease (PAD) affects tens of millions of Americans. It is a leading cause of lower extremity amputation. Health literacy is essential for disease awareness, but PAD awareness remains low, particularly in minoritized populations. The goal of the presented study is to investigate the PAD awareness and media preference for health information in Chinese-speaking communities in the metropolitans in California.

**METHODS:** An anonymous 14-question survey in Mandarin and English was designed to gauge basic knowledge of PAD, preferred methods for obtaining health information and general media preferences. The survey was distributed to health fairs in San Francisco (SF), Oakland, and Los Angeles (LA) in the Chinese-speaking communities. Associations between the above variables and demographics were compared between groups.

**RESULTS:** A total of 180 responses included 94 from SF, 57 from Oakland and 29 from LA. The SF cohort had a higher proportion of participants aged 65+ than Oakland (p = 0.018), however compared to the LA cohort, ages were similar. PAD awareness was low across all cohorts; however, diabetes was the most commonly identified factor affecting wound healing. LA and SF cohorts shared similar patterns to receive health information as they preferred radio (p = 0.021, SF 49/94 and LA 8/29) while the Oakland cohort favored video/media (p = 0.024, 18/57). For general media preferences SF and LA cohorts showed a stronger preference for newspapers (p = 0.471, SF 32/94 and LA 12/29) and television (p = 0.244, SF 28/94 and LA 12/29), while the Oakland cohort favored video (Oakland 20/57).

**CONCLUSION:** To our knowledge, this is the first study to survey multiple Chinese-speaking communities in the United States to assess PAD knowledge and learning preferences at the community level. The awareness of PAD among the selected Chinese-speaking communities was dismal. There was considerable variation in preferred learning methods between each selected community. Our findings underscore the importance of cultural and community-based health education strategies to improve PAD literacy.

**Human Ethics and Consent to Participate, approval Institutional Review Board (IRB) and funding declarations:** All participants were consented before proceeding to the survey. Nonetheless, the standard of the Declaration of Helsinki was followed in this study. The study is exempted and approved by Samuel Merritt University Institutional Review Board (SMUIRB #22-011). The presented study had no funding support.

## INTRODUCTION

Peripheral arterial disease (PAD) is a devastating and widely prevalent condition as it affects more than 12 million people in the United States. ^1^ Risk factors related to PAD include advanced age, diabetes, cigarette smoking and chronic kidney disease which are similar to other cardiovascular diseases. ^2^ It is a devastating condition due to its link to increased risk of limb loss, which is associated with increased mortality. The relationship between lower extremity amputation is thought to be due to the decline of cardiovascular function secondary to the loss of mobility.^3^ Lower extremity amputation in individuals with diabetic foot disease is associated with high long-term mortality, largely driven by the underlying burden of vascular disease and systemic complications.^4^ In addition to limb loss, other debilitating complications include ischemic pain as well as gangrene and/or non-healing wounds that predispose patients for infection and prolonged antibiotics.^5^

Unlike other cardiovascular diseases such as heart disease and stroke that had promising effects of enhancing health literacy, public education about PAD appears to be behind until recent years.^5,6^ To our knowledge the awareness of PAD remains to be inadequate in the Chinese-speaking population in the United States. ^7^ This was demonstrated in our recent findings in the San Francisco Chinatown survey which revealed a substantial knowledge gap regarding PAD and its complications.^7^ Low health literacy correlates with increased hospitalizations, emergency care usage, and decreased preventive service utilization.^8^ It also relates to challenges in interpreting health messages, poorer health status, higher mortality rates, and elevated healthcare costs.^9^ As a result, improving health literacy will likely improve health outcomes and reduce healthcare costs.

The factors that influence an individual’s health literacy can be multifactorial and complex. The access to health information appeared to be heterogenous. In a recent survey in California investigating the use of the internet for health information, only 40% of elderly reported to use the internet for health information. Social economic status and minority status were found to be disproportionately less inclined to use the internet for health information.^10^ However, the nuance related to health information access within a particular minority group which may have a wide range of social economic status is now well known. To our knowledge, there has been even less focus on the Chinese-speaking population as surveys and researches often negate such a widely diverse population. A nuanced understanding of how different Chinese-speaking communities access and process health information is essential to improving PAD education, early detection, and prevention efforts. Addressing these disparities can enhance patient engagement, ultimately reducing PAD-related complications and improving lower extremity outcomes in these high-risk populations. The presented study aimed to investigate PAD baseline knowledge and preferred media of choice to receive health information among Chinese-speaking populations in the US.

## METHODS

An anonymous 14-question survey which was tested in the prior study in Mandarin and English to gauge the basic knowledge of PAD and preferred method for obtaining health information.^7^ **[Supplemental A]**. The survey was distributed to Chinatown health fairs in San Francisco (SF) on May 6 and October 14, 2023, Oakland on July 14, 2024 and Los Angeles (LA) on November 9, 2024 targeted towards the Chinese-speaking communities. To our knowledge, there was no validated survey instrument in Chinese-language available; however, the survey that was used in the study was tested in our pilot study in San Francisco. For respondents who were not able to read the survey, trained student volunteers or certified translators read the questions and recorded responses on paper forms. Responses were subsequently transferred to Qualtric, where the data were stored. Informed consent to participate in the survey was acquired prior to the beginning of the survey although we did not collect any personal identifier. Human Ethics and Consent to Participate declarations - all participants were consented before proceeding to the survey. Nonetheless, the standard of the Declaration of Helsinki was followed in this study. The study is exempted and approved by Samuel Merritt University Institutional Review Board (SMUIRB #22-011). The presented study had no funding support.

### Variables

Key demographic variables collected included participants’ age, sex, education level, and English proficiency. Age was categorized into two groups (under 65 vs. 65 and older), consistent with previous literature linking advanced age to health decision-making. Education level was classified into high school or below and above high school to evaluate the impact of formal education on health literacy. English proficiency was grouped into proficient and limited proficiency categories, based on participants’ self-rated language skills. We categorized “proficient” for participants who reported “Extremely well”, “Very well” or “Well.” Respondents who reported “Not at all”, “Not too well” or “Not well” were under the “Limited proficiency” category. Survey consent was included in the first page.

### Statistical Analysis

The Chi-squared analyses were conducted to identify significant differences in demographics, media preferences, and learning styles between and within cohorts of the study. These analyses were conducted comparing each cohort to the San Francisco cohort, using it as a reference group to examine differences. This approach was chosen because our prior paper focused solely on the San Francisco cohort, making it a natural point of comparison, though we acknowledge that chi-squared tests do not establish a true control group.^7^ Additionally, chi-squared tests of independence were conducted to assess differences in the proportions of participants who selected each media type as a top choice for both general media preference and media used to receive health-related information. ANOVA analysis was also performed to investigate associations between preferred learning methods and both sex and education level. Logistic regression was conducted to further assess the top three preferred learning methods against English fluency, gender, age, and educational level. The significance level for all statistical tests were set at p-value of less than 0.05. All analyses were performed using Jeffreys’s Amazing Statistics Program (JASP; JASP Team 2024) and RStudio (RStudio Team 2024).

## RESULTS

Overall, a total of 180 responses included 94 from SF, 57 from Oakland, and 29 from LA. Sex distribution did not differ significantly between the SF cohort and other sites(Oakland: p = 0.506; LA: p = 0.630), with most participants self-identifying as female. In terms of age group, the SF cohort had a significantly higher proportion of participants aged 65 and older compared with Oakland (p = 0.018; SF: 78/92, Oakland: 39/57). English proficiency was similar between SF (26/91) and Oakland (22/55) but differed significantly from LA (2/25, p = 0.033). English proficiency differed significantly across these cohorts (p = 0.003; SF: 26/91, Oakland: 22/55, LA: 2/25), particularly among those with a high school education or below (p = 0.009; SF: 5/56, Oakland: 6/27, LA: 0/17). No significant differences in education attainment were observed between SF and the Oakland (p = 0.422) or LA (p = 0.433) cohorts. Primary language also differed significantly, with Chinese (including Cantonese, Mandarin and Toishanese) being the most commonly reported language in all cities (SF: 82/93; Oakland 40/57; LA 26/28). The demographic distribution is summarized in **Table 1**. Baseline knowledge of PAD and its risk factors was low across all cohorts. Specifically, no significant differences were found between the SF cohort and the others regarding prior awareness of PAD (SF: 32%; Oakland: 23%, p = 0.234; LA: 30%, p = 0.990)) or familiarity with PAD risk factors (SF: 25%; Oakland: 20%, p = 0.467; LA: 30%, p = 0.654). The total counts for individual demographic categories diddo not always sum to 180 due to variable item non-response, as participants may havecould skipped questions. Furthermore, the total count for primary language exceeds 180 as some respondents could sselected more than one language, and frequencies for PAD awareness reflect only the participants who responded affirmatively. Nonetheless, all respondents completed the entire survey.

**Table 1.** Surveyed Population Demographics.

|  |  | **SF (n=94)** | **OAKLAND (n=57)** | **LA (n=29)** | **Pooled (n=180)** |
| --- | --- | --- | --- | --- | --- |
| Sex | **Female** | 64 (68%) | 40 (70%) | 17 (59%) | 121 (67%) |
|  | **Male** | 27 (28%) | 13 (23%) | 9 (31%) | 49 (27%) |
|  | *p*-Value | - | *p* = 0.506 | *p* = 0.630 |  |
| Age | **65 year-old and older** | 78 (85%) | 39 (68%) | 24 (83%) | 141 (78%) |
|  | **younger than 65** | 14 (15%) | 18 (32%) | 5 (17%) | 37 (20%) |
|  | *p*-Value | - | *p* = 0.018 | - |  |
| Education Level | **College and above** | 30 (33%) | 19 (33%) | 5 (17%) | 54 (30%) |
|  | **High school** | 17 (19%) | 21 (37%) | 3 (10%) | 41 (23%) |
|  | **Below high school** | 43 (48%) | 12 (21%) | 15 (51%) | 70 (39%) |
|  | *p*-Value | - | - | - |  |
| PAD Awareness | **Had heard of PAD previously** | 29 (31%) | 13 (23%) | 8 (28%) | 50 (28%) |
|  | *p*-Value | - | *p* = 0.234 | *p* = 0.990 |  |
|  | **Familiarity with PAD risk factors** | 22 (23%) | 11 (19%) | 8 (28%) | 41 (23%) |
|  | *p*-Value | - | *p* = 0.467 | *p* = 0.654 |  |
| English Proficiency | **Proficient** | 25 (28%) | 22 (39%) | 2 (7%) | 49 (27%) |
|  | **Limited proficiency** | 65 (72%) | 33 (58%) | 23 (79%) | 121 (67%) |
|  | *p*-value | - | *p* = 0.154 | *p* = 0.033 |  |
| Primary Language | **Chinese** | 82 (89%) | 40 (73%) | 26 (96%) | 148 (82%) |
|  | **English** | 15 (16%) | 20 (36%) | 2 (7%) | 37 (20%) |
|  | *p*-value | - | *p* = 0.016 | *p* = 0.413 |  |

**Note:** Total counts for single-choice categories (Sex, Age, Education, and English Proficiency) may be less than 180 due to missing or skipped responses. The total for Primary Language exceeds 180 because respondents may have selected multiple languages. PAD awareness counts reflect only those who answered affirmatively.

### Preferred Method(s) to Receive Health Information

Significant differences were observed in preferred learning modalities across cohorts. SF participants were more likely to report radio as a top method of health information compared with Oakland (p < 0.001; SF: 49/94, Oakland: 11/57) and LA (p = 0.021; SF: 49/94, LA: 8/29).. Preferences for video/media also differed between SF and Oakland (p = 0.024; SF: 15/94, Oakland: 18/57) and SF also differing (p = 0.028; SF: 15/94), but not between SF and LA (p = 0.870). Internet-based health information sources were not significantly different between SF and Oakland (p = 0.598) and SF and LA (p = 0.870). No significant differences were found across cohorts in preferences for email-based health information reception (Oakland: p = 0.254; LA: p = 0.553).

### Preferred General Media Sources

Regarding newspapers as a preferred media source, SF and Oakland showed a trending difference (p = 0.089; SF: 32/94, Oakland: 12/57), while SF and LA (p = 0.471) were similar. For video/media as a preferred media source, SF and Oakland differed significantly (p = 0.007; SF: 15/94, Oakland: 20/57), as did SF (p < 0.001; SF: 15/94). No significant difference was found between SF and LA (p = 0.315). No significant differences were found for television as a preferred media source across any of the cohorts (Oakland: p = 0.265; LA: p = 0.244). For Apps as a preferred media source, SF differed significantly from LA (p = 0.035; SF: 13/94, LA: 9/29). SF and Oakland showed a trending difference (p = 0.095; SF: 13/94, Oakland: 14/57).

### Media Preferences for Health Information

Across SF, LA and Oakland cohorts, men had different proportions of using video/media to learn about healthcare (p = 0.018; SF: 1/26, Oakland: 5/8, LA: 2/7) compared to women (SF: 14/50, Oakland: 13/27, LA: 3/14). Additionally, among individuals aged 65 and older, the proportion who used video/media as their primary media type differed significantly (p = 0.026; SF: 13/65, Oakland: 10/29, LA: 5/19). Media preference was also associated with English proficiency and education level, with participants who had higher educational attainment and greater English proficiency more likely to prefer written or digital formats, such as email or internet-based learning. Among Oakland participants specifically, the proportion of individuals who reported video/media as a preferred media source differed significantly by age group (p = 0.028; <65: 10/8, 65 and up: 10/29), with younger participants showing a higher preference (56%) compared to those 65 and older (26%). Additionally, preference in using Chat apps as a preferred media source also varied significantly by age within the Oakland cohort (p = 0.018; <65: 10/8, 65 and up: 33/6). For participants from the LA cohort, education level was significantly associated with whether individuals considered email as a top method to receive health information (p = 0.005; >HS: 2/3, HS and below: 0/18).

Among all media types, only radio and video were endorsed by participants in both contexts; all other media types were either absent from one category entirely or reported too infrequently to allow for analysis. For radio, no significant association was found between general and health-specific media selection, either in the combined sample or within individual cohorts (p > 0.05), indicating that participants’ responses to the two questions were statistically independent — selecting radio in one context did not make them more or less likely to select it in the other. In contrast, for video, a significant association was observed both in the overall cohort (p < 0.001; 57.1% who selected video as a general media preference also selected it as a preferred source of health information) and within each individual cohort (SF: p < 0.001; 60%, Oakland: p = 0.005; 55%). This indicates that participants who selected video as a top general media preference were significantly more likely to also select it as a preferred source for health-related information, and vice versa, suggesting a non-random, bi-directional pattern of endorsement across the two contexts.

## DISCUSSION

To our knowledge, this was the first attempt to investigate the baseline PAD knowledge and preferred method to acquire health information of a subset of the Chinese-speaking population across multiple metropolitans in California, United States. In the presented study, we expanded our previous work to include additional Chinese-speaking communities including Oakland and LA.^7^ A number of interesting findings were identified as compared to the SF cohort. The lack of awareness of peripheral arterial disease (Oakland, p= 0.234; LA, p= 0.990) and its risk factors (Oakland, p= 0.467; LA, p= 0.654) regardless of the educational level and age distribution of the community. The variation in preferred learning methods from community to community as identified in the findings also highlighted potential challenges on how we may improve the PAD awareness in these communities.

San Francisco, which served as the reference for comparison with newly surveyed communities.^7^ The SF cohort which was characterized by an older age distribution (78/92, 85%) and moderate education levels (43/90 below high school, 48%), with low English proficiency (65/90, 72%) had a strong preference to learn health information via traditional channels such as radio (49/94, 52%). The cohort also preferred newspapers (32/94, 34%) as the media source in general. When examining all surveyed sites together, a pattern of significant variation in learning preferences emerged, associated not only by location but by underlying demographic factors. As suggested by the literature, life experiences and generational background can impact adulting learning.^11^ This was reflected in our findings, where older participants, particularly in SF (78/92, 85%) and LA (24/29, 83%) which have a significantly higher proportion of individuals aged 65 and older, were more likely to prefer radio (SF 49/94, 52% and LA 8/29, 28%) compared to their counterparts in Oakland (11/57, 19%). Meanwhile, participants in Oakland who were 65 years old and younger favored video/media (32%) or internet-based learning (19%). These preferences underscored how generational background may be related to not only comfort with different media formats but also the effectiveness of educational outreach. **[Figure 1-3, Table 2]** Therefore, no single media may suit all Chinese-speaking communities to improve health literacy. Although PAD awareness itself remained uniformly poor across all surveyed sites, the way information was accessed and understood differed remarkedly. This emphasized the culturally similar populations may require distinctly localized and demographically aligned outreach strategies to effectively improve health literacy.

**Table 2.** Preferred Methods to Receive Health Information in comparison to SF cohort.

|  |  | SF<br>(n=94) | OAKLAND<br>(n=57) | LA<br>(n=29) |
| --- | --- | --- | --- | --- |
| Radio |  | 49 (52%) | 11 (19%) | 8 (28%) |
| | *p*-Value | - | $p < 0.001$ | $p = 0.021$ |
| Video/Media |  | 15 (16%) | 18 (32%) | 5 (17%) |
| | *p*-Value | - | $p = 0.024$ | $p = 0.870$ |
| Internet-based learning |  | 15 (16%) | 11 (19%) | 5 (17%) |
| | *p*-Value | - | $p = 0.598$ | $p = 0.870$ |

**Figure 1.**
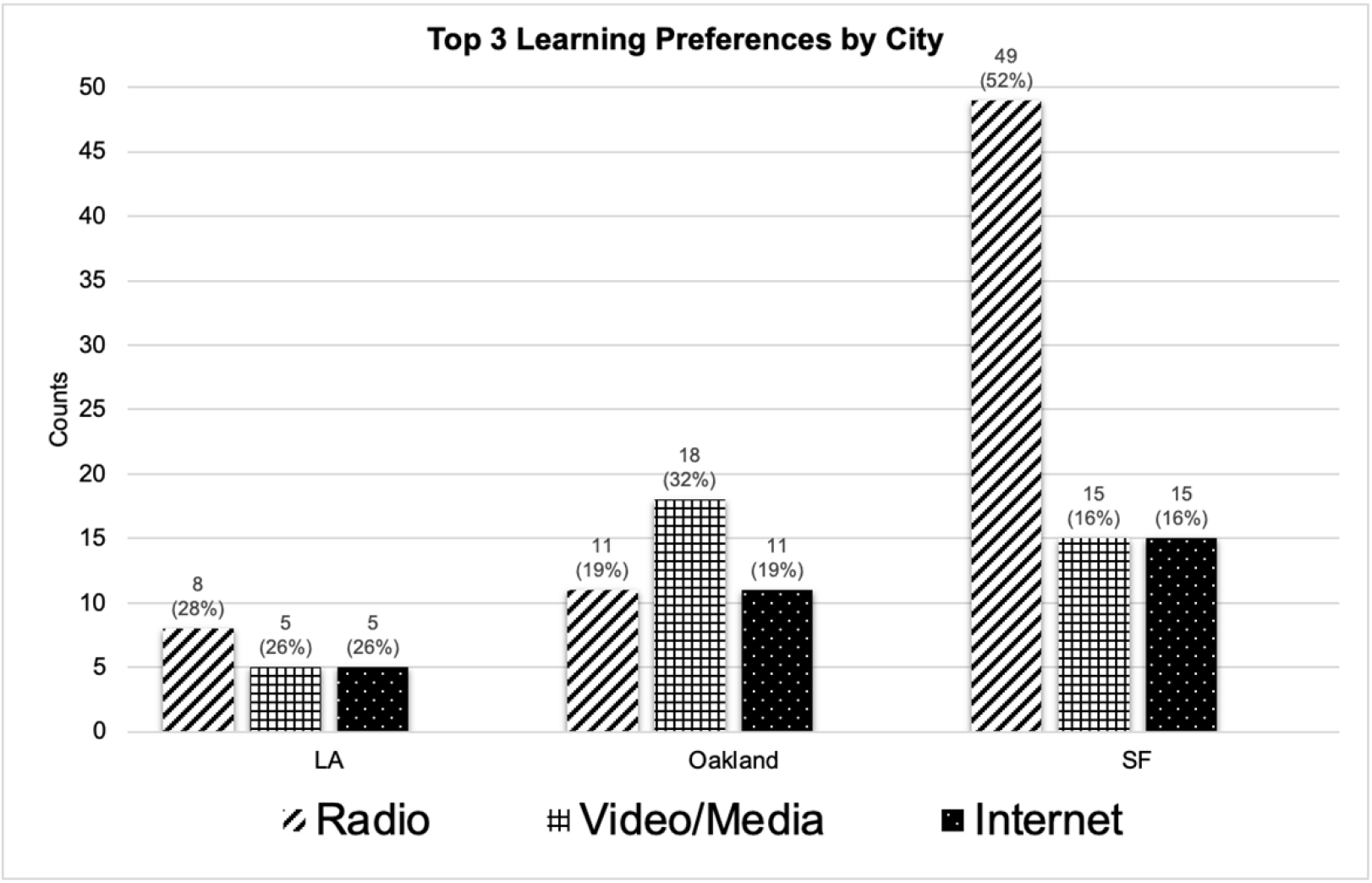
Number of respondents in each city who selected various learning methods among their top three. Radio was the most common in SF, while Oakland favored video/media.

**Figure 2.**
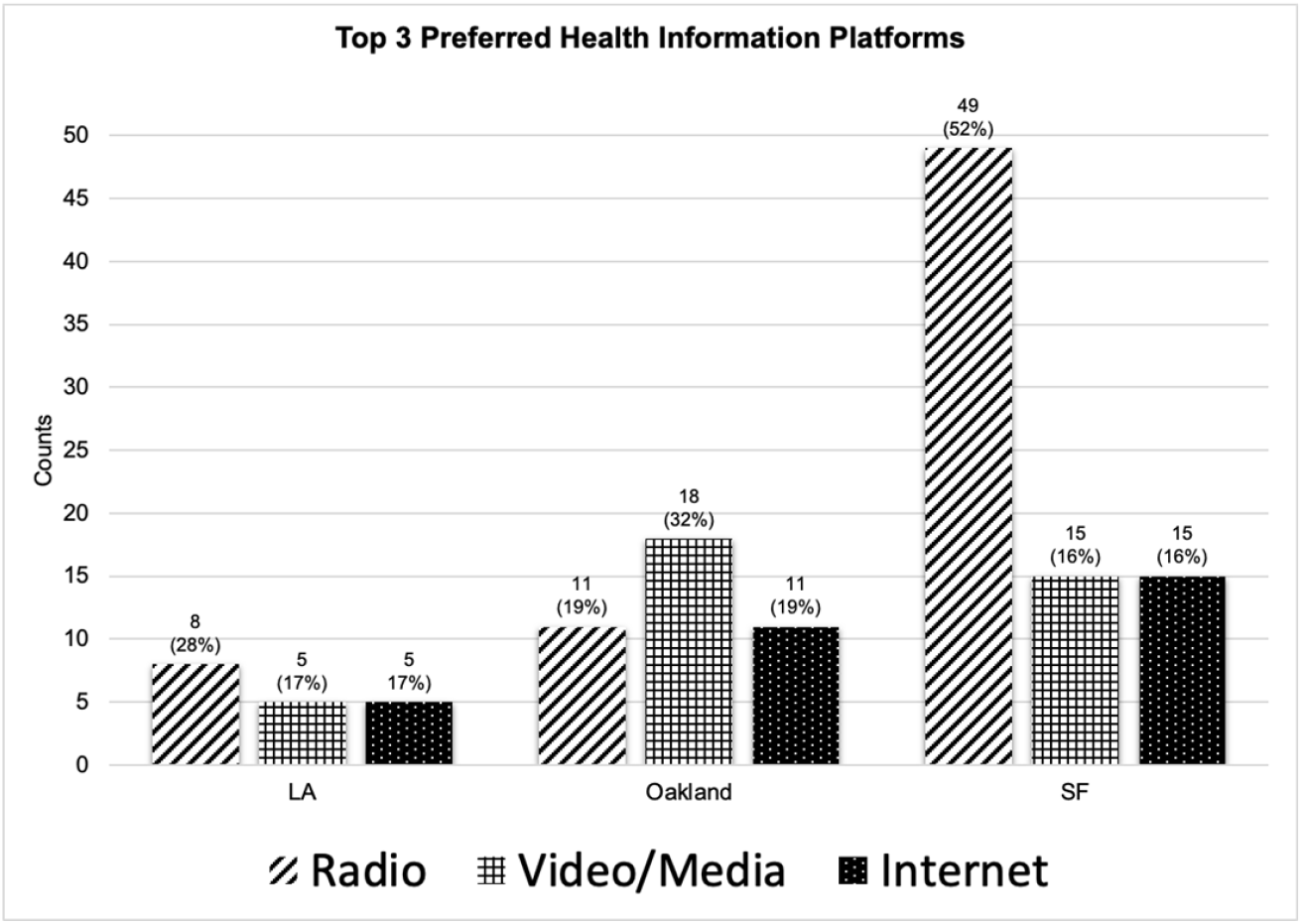
Number of respondents in each city who selected various methods for receiving health information among their top three. Radio was the most common in SF, while Oakland favored video/media.

**Figure 3.**
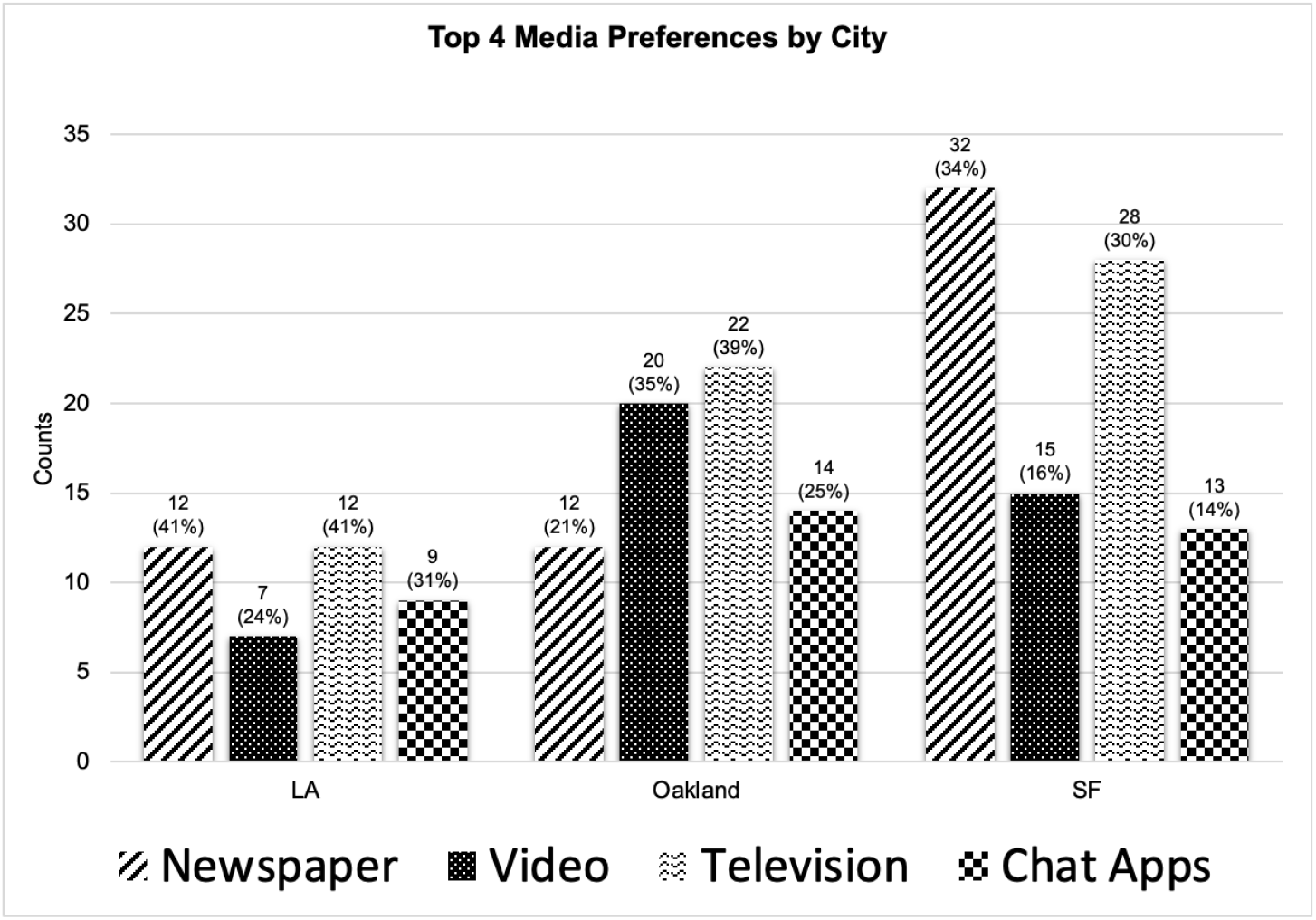
Number of respondents in each city who selected various general media sources among their top four. SF and LA showed a stronger preference for newspaper and television, while Oakland favored video as their media preference.

Findings of our cohorts suggested that the awareness of PAD was far behind the awareness of other cardiovascular diseases. According to the National Health Interview Survey (NHIS) between 2009 to 2014, greater than 90% respondents of a survey recognized that sudden numbness of one side of the body is a sign of stroke and 68.3% of respondents recognized all 5 symptoms.^12^ The authors concluded that stroke awareness was improving but still not optimal. Similarly, the awareness of heart attack is much improved in the similar survey.^13^ However, the author indicated the influence of socioeconomic status implicated the disparity in the knowledge and multifaceted approach would be necessary to improve the awareness. The above findings related to stroke and heart attack coincided with our results, although the awareness of peripheral arterial disease, such as having heard of PAD (SF 29/91, 32%; Oakland 13/57, 23%; LA 8/27, 30%) and knowing its risk factors (SF 22/87, 25%; Oakland 11/55, 20%; LA 8/27, 30%) was much more dismal across our survey communities than stroke and heart attack, even though some respondents and communities had relatively high education. The successful improvement of stroke and heart attack awareness in the general population was largely driven by highly visible, multifaceted public health campaigns utilizing diverse media formats, from television broadcasts to targeted digital media. In contrast, PAD lacks this level of systemic, multimedia public health messaging. While the cause of such low awareness may be multifactorial.

When comparing our findings to health literacy studies in other populations, our results align with existing evidence highlighting a reliance on traditional and ethnic media among older and minoritized groups. For example, studies examining health education in Hispanic/Latino and African American populations consistently show that older adults heavily rely on culturally specific radio, television, and print media to receive trusted health information.^14^ This mirrors our older SF and LA cohorts, who demonstrated a strong preference for radio and newspapers. Conversely, the Oakland cohort’s preference for video and internet-based learning aligns with broader national trends showing a shift towards digital health information-seeking among younger or higher-educated demographics.^15^ This phenomenon reflects the well-documented “digital divide” in health literacy, where age, education, and language proficiency dictate whether a population prefers a traditional community broadcasts over digital modalities.^16^

As witnessed in heart attack and stroke campaigns, improving awareness benefits from a multifaceted approach. Furthermore, our project suggested that to replicate this success for a PAD awareness campaign, the approach must be tailored to individual communities. Even within a single ethnic group sharing a common cultural background and primary language, we observed distinct variation in preferred learning methods. Specifically, prior study indicated that higher educational attainment was positively correlated with electronic health literacy.^17^ In our survey, this correlation was similarly observed, as participants with education beyond high school were more inclined toward digital and written learning formats. For example, in the LA cohort, educational level was significantly associated with a preference for email as a top method to receive health information (p = 0.005), suggesting that both age and educational level were associated with both the capacity to understand health information but also the mode through which individuals are most receptive to receiving it. Language, too, plays a crucial role in shaping health communication effectiveness. Despite statistically significant differences across cities, Chinese languages - including Cantonese, Mandarin, and Toishanese on the survey - remained the predominant primary language among respondents in this study (SF 89%; Oakland 73%; LA 96%). These findings support the development of linguistically appropriate interventions that prioritize Chinese-language materials while also accommodating bilingual resources where needed. Low health literacy was shown to correlate with increased hospitalizations and emergency care usage which attribute higher health care cost let alone the challenge to interpret health messages.^8^ As a result, people with low health literacy were more likely to have poor health outcomes including higher mortality rate.^9^

The above findings underscored the importance of moving beyond a uniform outreach strategy and instead adopting a community-specific framework for improving PAD awareness. By identifying how demographic characteristics shape both health literacy and communication preferences, future interventions can be more intentionally designed. In some communities, this may mean utilizing radio broadcasts for older, less digitally connected populations or leveraging digital media for younger, more educated audiences. Partnering with trusted local organizations will also be critical in disseminating tailored education. Moreover, these insights lay the groundwork for developing multi-lingual, multimedia educational tools and for expanding similar assessments to other underrepresented groups. Ultimately, translating this knowledge into targeted culturally sensitive action is essential for closing the gap in PAD awareness and reducing disparities in vascular health outcomes.

## Limitations

The current findings have some limitations. First, the sampling of the population took place during health fairs which could be subject to bias for the subpopulation that could be more health conscious. Additionally, we did not track refusal rates or the demographics of non-participants, and some respondents skipped certain questions. Although this may raise concern of potential selection bias, respondents went through all the survey questions and we analyzed all collected responses. However, the awareness of such subpopulation had very low knowledge about PAD indicating that the awareness of PAD for the remaining community may be similar or more dismal. Second, due to limited resources, we were not able to sample cities and other Chinese-speaking communities outside of Chinatown. Under such constraints, we also could not survey more people in each event. Nonetheless, the survey process was standardized across all surveyed sites to mitigate potential bias. Like many survey studies, all data were self-reported, which is subject to recall bias. The study employed a cross-sectional design, which limits the ability to infer causality or track changes in knowledge over time. Lastly, unmeasured confounders such as country of origin and years of residence in the U.S. may have influenced both awareness and media preferences and were not controlled for in the analysis. This comparison between general media preference and health-related media use was limited to radio and video, as other media types lacked sufficient overlap across both contexts. As a result, the findings may not generalize to broader media patterns, and other media types may show different associations that could not be explored in this analysis. We hoped that the presented study would lay the foundation for a larger effort to include more Chinese-speaking communities as well as other minority groups so the awareness of PAD can be in part with other cardiovascular diseases.

## CONCLUSION

To our knowledge, this was the first attempt to survey multiple Chinese-speaking communities in the United States to gauge the knowledge of PAD and to understand learning method preferences in different communities. The findings suggest the need to improve awareness and knowledge of PAD. Although each Chinese-speaking community shares a similar cultural background, the social determinants vary from one community to another. As a result, the strategy to improve such awareness in each community should be individualized and multifaceted. The presented work provided a framework to understand other minority communities beyond Chinese-speaking ones in the future as our findings suggested nuances within a specific ethnic group. Similar nuances may exist beyond Chinese-speaking communities. Considering the recent trend of personalized medicine, prevention should also be precise and individualized to be most effective.

## Data Availability

Raw data from this study is made available in the Supplementary file.

## LIST OF ABBREVIATION

LA: Los Angeles
PAD: Peripheral arterial disease
SF: San Francisco

